# Baseline phenotype and 30-day outcomes of people tested for COVID-19: an international network cohort including >3.32 million people tested with real-time PCR and >219,000 tested positive for SARS-CoV-2 in South Korea, Spain and the United States

**DOI:** 10.1101/2020.10.25.20218875

**Authors:** Asieh Golozar, Lana YH Lai, Anthony G. Sena, David Vizcaya, Lisa M. Schilling, Vojtech Huser, Fredrik Nyberg, Scott L. Duvall, Daniel R. Morales, Thamir M Alshammari, Hamed Abedtash, Waheed-Ul-Rahman Ahmed, Osaid Alser, Heba Alghoul, Ying Zhang, Mengchun Gong, Yin Guan, Carlos Areia, Jitendra Jonnagaddala, Karishma Shah, Jennifer C.E. Lane, Albert Prats-Uribe, Jose D. Posada, Nigam H. Shah, Vignesh Subbian, Lin Zhang, Maria Tereza Fernandes Abrahão, Peter R. Rijnbeek, Seng Chan You, Paula Casajust, Elena Roel, Martina Recalde, Sergio Fernández-Bertolín, Alan Andryc, Jason A. Thomas, Adam B. Wilcox, Stephen Fortin, Clair Blacketer, Frank DeFalco, Karthik Natarajan, Thomas Falconer, Matthew Spotnitz, Anna Ostropolets, George Hripcsak, Marc Suchard, Kristine E. Lynch, Michael E. Matheny, Andrew Williams, Christian Reich, Talita Duarte-Salles, Kristin Kostka, Patrick B. Ryan, Daniel Prieto-Alhambra

## Abstract

Early identification of symptoms and comorbidities most predictive of COVID-19 is critical to identify infection, guide policies to effectively contain the pandemic, and improve health systems’ response. Here, we characterised socio-demographics and comorbidity in 3,316,107persons tested and 219,072 persons tested positive for SARS-CoV-2 since January 2020, and their key health outcomes in the month following the first positive test. Routine care data from primary care electronic health records (EHR) from Spain, hospital EHR from the United States (US), and claims data from South Korea and the US were used. The majority of study participants were women aged 18-65 years old. Positive/tested ratio varied greatly geographically (2.2:100 to 31.2:100) and over time (from 50:100 in February-April to 6.8:100 in May-June). Fever, cough and dyspnoea were the most common symptoms at presentation. Between 4%-38% required admission and 1-10.5% died within a month from their first positive test. Observed disparity in testing practices led to variable baseline characteristics and outcomes, both nationally (US) and internationally. Our findings highlight the importance of large scale characterization of COVID-19 international cohorts to inform planning and resource allocation including testing as countries face a second wave.

## INTRODUCTION

As of 22^nd^ October 2020, more than 41 million confirmed cases of coronavirus disease 2019 (COVID-19), and more than 1.1 million deaths have been reported worldwide^1^. As many countries emerge from the impact of the initial phase of the COVID-19 pandemic, strategies to contain the virus are rooted in the rapid testing and identification (‘test and trace’), and subsequent isolation (quarantine) of emerging cases to mitigate community transmission and future outbreaks^2^.

Great heterogeneity exists in testing policies worldwide, with some countries performing population-scale mass testing, and others opting for more selective testing approaches. Testing availability has also changed rapidly over time. Initially, testing in many countries was limited to only severe cases or those with symptoms, but that has rapidly changed as resources and testing capabilities have become more widely available^3^. Spain, for example, had most of the testing carried out in hospitals in the initial phase of the pandemic, but has now implemented primary care and community mass screening. However, it is likely that testing capacity will be insufficient again as second waves of COVID-19 arise in different countries. This is becoming obvious in the United Kingdom (UK) and the United States (US) amongst others, where tests are sometimes unavailable and/or delayed beyond what is needed for effective test-tracing^4, 5^.

Although aggregated figures are updated regularly and in the public domain, there is a scarcity of linked patient-level data on the clinical characteristics and outcomes of those tested and those tested positive^1, 6^. While many previous reports have described people tested positive or with a confirmed diagnosis of COVID-19, only a minority focused on population-based cohorts. In addition, most of such literature consists of case reports or case series, with only a very small minority including at least 1,000 participants.

Understanding the characteristics at the time of testing and health outcomes following a positive test is crucial, not only to understand disease severity and spectrum of illness, but also to provide information for forecasting COVID-19 spread and healthcare provision in the coming months. In this study, we aimed to characterize baseline socio-demographics, comorbidities and symptoms seen at the time of testing, and to assess key health outcomes amongst those tested and those tested positive.

## METHODS

### Study Design

We performed a large-scale cohort study across a network of outpatient and inpatient electronic health records (EHRs) and national claims data, all standardized to the Observational Medical Outcomes Partnership (OMOP) Common Data Model (CDM)^7^. The OMOP CDM and associated tools were used to conduct large scale federated analytics within the Observational Health Data Sciences and Informatics (OHDSI) global research network, whereby each participating institution retained their patient-level data, and run analytical code made available to them by analysts for remote querying, i.e. in a distributed network fashion^8^. This study is part of the “Characterizing Health Associated Risks, and Your Baseline Disease In SARS-COV-2 (CHARYBDIS)” study. The study protocol is available at https://github.com/ohdsi-studies/Covid19CharacterizationCharybdis/blob/master/documents/Protocol_COVID-19%20Charybdis%20Characterisation_V5.docx

### Data

Of twenty-one databases contributing data to OHDSI’s open science community efforts to characterize the natural history of COVID-19, 12 were included in this study. Nine data sources were excluded due to absence of testing data, insufficient (<1 year) follow-up before testing or where test data were unavailable. Included data in the study were obtained from US health systems including Columbia University Irving Medical Center (CUIMC) in New York (NY), STAnford medicine Research data Repository (STARR-OMOP) in California (CA)^9^, Tufts CLARET (TRDW) in Massachusetts (MA), University of Colorado Anschuz Medical Campus Health Data Compass (CU-AMC HDC) in Colorado (CO) and UW Medicine COVID Research Dataset (UWM-CRD) in Washington (WA) state; US-wide hospital data from Premier, Optum® de-identified COVID-19 Electronic Health Record dataset (Optum EHR), and the Department of Veterans Affairs (VA-OMOP)^10^; and national claims data from IQVIA Open Claims and HealthVerity. Data from South Korea (SK) came from nation-wide claims recorded in the Health Insurance Review & Assessment (HIRA) database. Spanish data came from the Information System for Research in Primary Care (SIDIAP), a primary care database from Catalonia linked to hospital admissions and regional COVID-19 tests. Data collection period varies by database from January 2020 to the end of data collection. Most data were collected from March to May 2020, whilst the data collection period for five databases span to June 2020 and beyond (CUIMC, HealthVerity, STARR-OMOP, UWM-CRD and VA-OMOP). Details on the included data sources are available in Supplementary Figure 1 and Supplementary Tables 1 and 2.

### Study Participants

Two cohorts were studied: 1) individuals with a first real-time PCR SARS-CoV-2 test performed between January 2020 and June 2020 (*tested* cohort), 2) individuals who tested positive for SARS-CoV-2 for the first time during the same timeframe (*tested*+ cohort). The respective index dates were the day of the first test for the *tested* cohort, and the test sample collection date of the first positive test for the *tested*+ cohort. Cohort participants were followed from index date to the earliest of death, end of the study period, or 30 days after index. The codes used to identify COVID-19 diagnoses and SARS-CoV-2 test are described in more detail in Supplementary Table 3. All study participants were required to have at least 365 days of prior observation time before index date to allow comprehensive capture of comorbidities and medication use.

Having a negative test for SARS-CoV-2 is a transient status and dependent on the timeframe of the study. A person tested negative for SARS-CoV-2 could test positive for SARS-CoV-2 later in time and be considered a part of the tested positive cohort if the new test was performed during the timeframe of the study. This particularly makes it difficult to study this cohort during the pandemic. As such, we did not report on the symptoms and characteristics of persons tested negative for SARS-CoV-2 test.

### Baseline characteristics and outcomes of interest

Demographic data including age, sex and symptoms at index date were obtained. Comorbidities from day -30 to day -1 before index date were identified based on the SNOMED CT hierarchy, with all descendant codes included. The key conditions reported here are based on recent reports of associations with COVID-19 infection or outcomes^11^. All conditions are reported in full at an interactive <u>website</u>, although this is dynamic and will change as new participants and/or databases are added over time. Detailed definitions of each condition are available at http://atlas-covid19.ohdsi.org/#/home.

The main study outcomes were ascertained from index to day 30 post-index and included hospitalization and death. Additional respiratory, cardiovascular (i.e. acute myocardial infarction, sudden cardiac death, ischemic stroke, intracranial bleed and heart failure), Venous thromboembolism events (i.e. pulmonary embolism and deep vein thrombosis) and other secondary outcomes were also analyzed.

### Statistical analyses

All features and outcomes were summarized as proportions and reported per database separately. Databases were grouped into primary care EHR, hospital (inpatient and outpatient) EHR, and claims for reporting and plotting purposes to highlight the differences in the nature of the source of data.

All analytical code was developed centrally and run locally in each database in a federated manner, and is available at https://github.com/ohdsi-studies/Covid19CharacterizationCharybdis. The data reported here were extracted from the CHARYBDIS results set (available <u>here</u> in full) on 04/10/2020.

All the data partners obtained Institutional Review Board (IRB) approval or exemption, as appropriate, to conduct this study. Co-authors with access to the relevant documentation and with direct access to each of the contributing data sources are mentioned in Supplementary Table 1.

## RESULTS

A total of 3,316,107 persons were tested and 219,072 tested positive in the participating databases between January and June 2020 and were therefore included in the *tested* and *tested*+ cohorts respectively. Twelve databases (one primary care, eight hospital EHR, and three health claims databases) from three countries (Spain, SK and the US) were included (Supplementary Figure 1). For databases with both testing and test result data available (eight out of twelve), the ratio of positive/tested varied dramatically, from 2.3:100 in STARR-OMOP, 6.2 in VA-OMOP, 13.4:100 in HealthVerity, 22.5:100 in SIDIAP, and up to 31.2:100 in CUIMC. This ratio decreased over time, with much higher positive/tested ratios in February-April (50:100 in April) than in May-June (6.8:100 in June).

### Baseline demographics, comorbidities, and symptoms

The majority of study participants were aged 18-64 years old, with small representation of children and a variable proportion of elderly (>65) ranging from 20% to 48% of the total tested. Women were overrepresented amongst the tested (52% in HIRA to 64% in SIDIAP) in all data sources except VA-OMOP, which is a predominantly male cohort. Detailed patient characteristics including socio-demographics, pre-existing comorbidity and prior drug use are reported in Table 1.

**Table 1:**
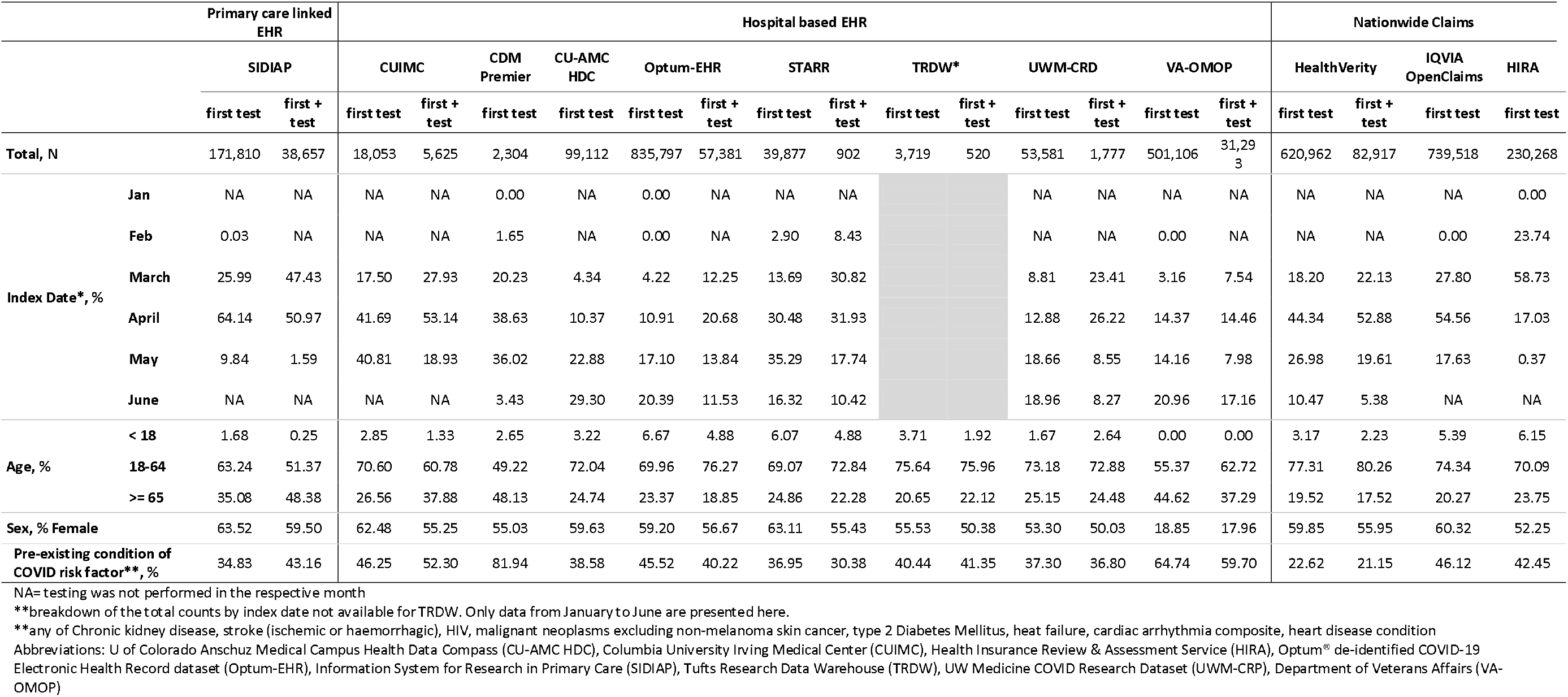
Characteristics of people tested with real-time PCR and patients tested positive for SARS-CoV-2 in a network of databases from Spain, South Korea and United States.

Key comorbidities were generally more prevalent in the tested+ compared to the overall tested (Figure 1, and age- and sex-stratified in Supplementary Figures 2). These differences were consistent across databases for hypertension (59.9% in the tested+ vs 20.1% in the tested in VA-OMOP), obesity (44.4% vs 31.2% in VA_OMOP) and heart disease (42.4% vs 18.8% in VA-OMOP).

**Figure 1:**
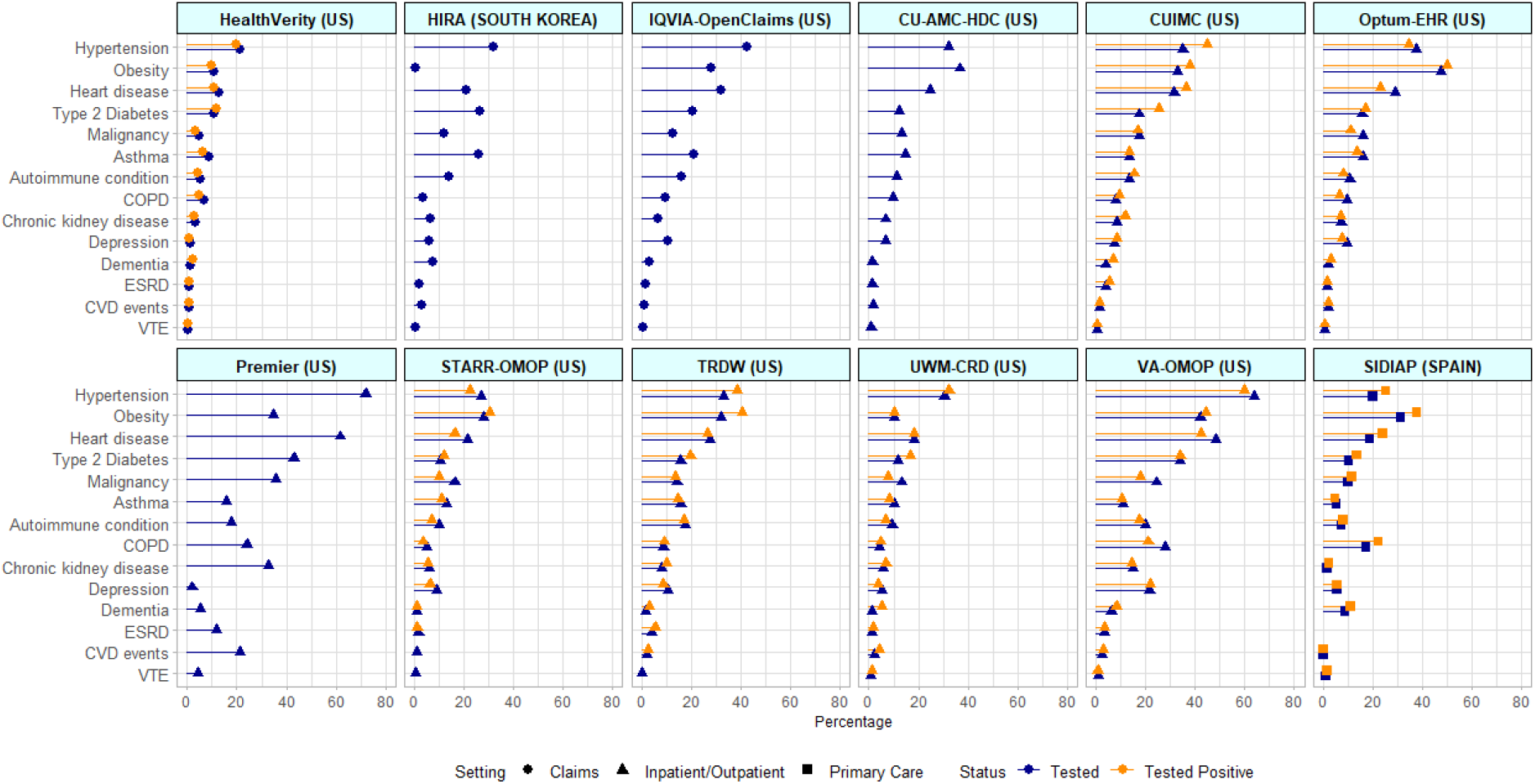
Baseline comorbidities 30-days prior to index date among SARS-CoV-2 *tested* and *tested*+ cohorts across databases of various setting. COPD = Chronic obstructive pulmonary disease; ESRD = End stage renal disease; CVD = Cardiovascular disease; VTE = Venous thromboembolism events; US= United States

The prevalence of symptoms at index date in those tested and tested+ is depicted in Figure 2 (stratified by age and sex in Supplementary Figures 3). The most commonly reported symptoms amongst the tested were cough (2.6% in UWM-CRD to 27.5% in IQVIA-OpenClaims), fever (from 1.1% in SIDIAP to 18.6% in HIRA), and dyspnoea (1.3% in SIDIAP to 15.1% in TRDW). The prevalence of these symptoms was higher in those tested+ and in hospital EHR compared to claims or primary care databases.

**Figure 2:**
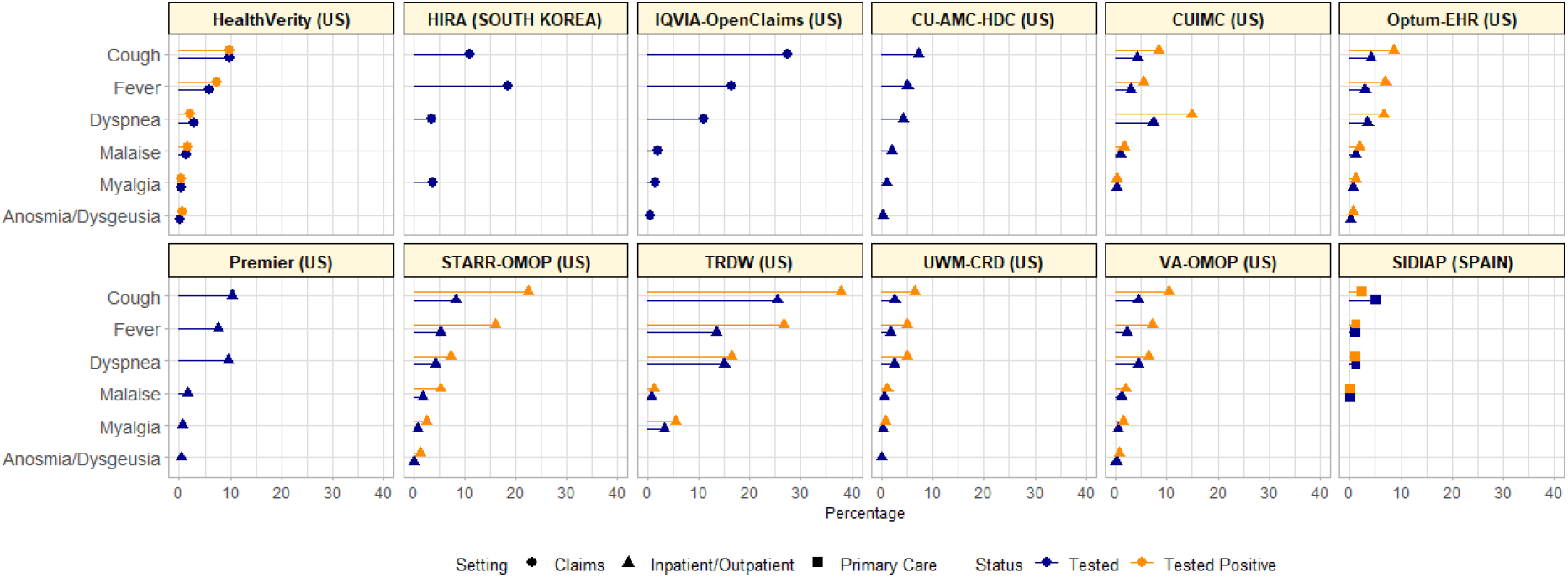
COVID-19 symptoms at index date among SARS-CoV-2 *tested* and *tested+* cohorts across databases of various setting.

### 30-day outcomes amongst the tested+

The percentage of patients hospitalized within 30 days after testing positive was: 4.2% (HealthVerity), 9.6% (STARR-OMOP), 19.5% (UWM-CRD), 19.8% (Optum-EHR), 22.9% (TRDW), 25.5% (VA-OMOP), 27.1% (SIDIAP), and 37.6% (CUIMC). Overall fatality in the month after the first positive test ranged from 9.2% (CUIMC) to 10.5% (SIDIAP) (Figure 3). There was a general downward trend over time in outcomes. For example, from March to May, hospitalization decreased from 45% to 14.3% and 30-day mortality decreased from 11% to 1.2% in CUIMC.

**Figure 3:**
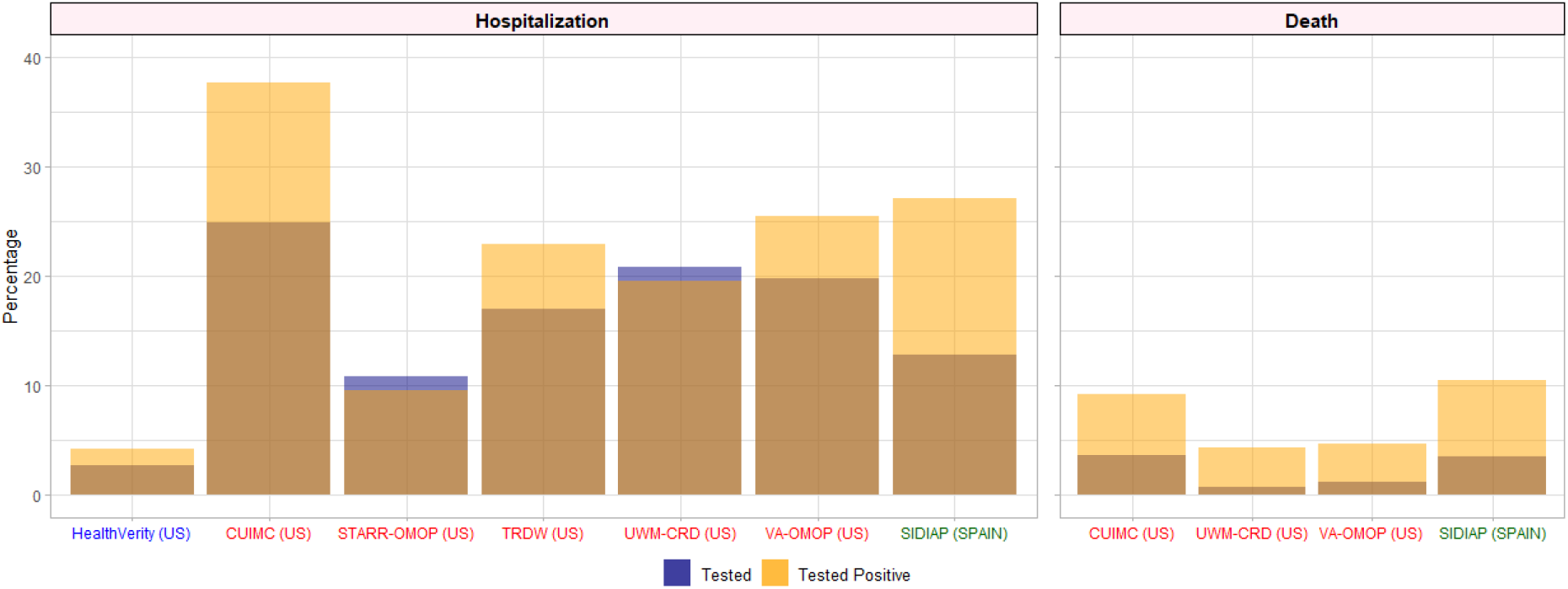
Hospitalization and death 30 days after first test or first positive test, respectively, among SARS-CoV-2 *tested* and *tested+* cohorts across databases of various setting. The overlapped area (brown) indicates an overlap between both tested and tested+ cohorts. Different coloured fonts indicate settings. Blue = Claims; Red =Hospital EHR; Dark green = Primary care HER

Regarding other complications, the percentage of *tested*+ patients diagnosed with pneumonia within 30 days from the test date ranged from 3.6% (HealthVerity) to 22.4% (VA-OMOP); acute respiratory distress syndrome (ARDS) from 1.1% (HealthVerity) to 12.0% (VA-OMOP); sepsis 0.6% (HealthVerity) to 4.9% (VA-OMOP). Acute kidney injury was the most common renal outcome (0.6% in HealthVerity to 7.9% in VA-OMOP), followed by dialysis (0.2% in HealthVerity to 1.5% in both TRDW and VA-OMOP). The composite of total cardiovascular events occurred in 0.2% (SIDIAP) to 5.0% (VA-OMOP), whilst venous thromboembolic events were recorded in 0.2% (HealthVerity) to 2.3% (VA-OMOP) (Supplementary Figure 4).

## DISCUSSION

Including over 3.32 million tested and over 219,000 tested positive, we report on the largest patient-level cohort to date of individuals tested for SARS-CoV-2 with linked baseline characteristics and health outcomes. We conducted our analyses in 12 real world data sources from three continents. Positive test ratios varied widely across geography from 2:100 in California to >25:100 in Spain and >30:100 in NYC, illustrating great heterogeneity in testing practice and extent of testing. Similar variability was observed over time in the first half of 2020, with much higher positive/test ratios in February-March, and lower in May-June.

Notwithstanding the known effect of age and male sex on COVID-19 severity, we show that a majority of those tested and tested+ were in fact adult (18-65 years) women. This is consistent with official sources from the US^12^, Spain^13^, and SK^14^. Plausible explanations include higher work-related exposure (i.e. hospital staff, care homes)^15-17^ and/or differences in healthcare seeking behaviour leading to increased testing in women^18^.

The most common comorbidities reported in previous systematic reviews were hypertension, cardiovascular disease and diabetes^19, 20^. In line with previous knowledge, hypertension, obesity and cardiovascular disease were the most prevalent comorbidities in the *tested*+ cohort with over 34% of the population suffering from at least one of these three conditions in the US^21^, which was consistent with our findings. The higher proportion of these chronic comorbid conditions can be due to the targeted testing of affected people, or a potential association between these conditions (or related treatments) and an increased susceptibility to SARS-CoV-2 infection^22, 23^.

The identified range of symptoms is wide, and coding differs across settings, with more symptoms being recorded in hospital data sources than primary care data such as SIDIAP. Despite this variability, there is a consistent recording of fever, cough and dyspnoea as the most common presenting symptoms at the time of testing. All of these symptoms are more common amongst the *tested*+ than in the overall tested population. Despite under-reporting of symptoms in our study, our findings are consistent with other published literature, including reports from the Centers for Disease Control and Prevention (CDC)^24^ and the World Health Organization (WHO)^25^. This lower prevalence compared to research cohorts may be due to incomplete capture of symptoms in coded/structured records. More work is needed to further unravel the potential discriminatory ability of these symptoms, alone or combined, for the differential diagnosis of COVID-19 in the presence of competing influenza and other viruses causing influenza-like disease.

Finally, we studied a number of health outcomes associated with COVID-19 infection. Overall, 4% to 38% of the tested+ cohort required hospitalization, and 4% to 11% died within a month of their first positive test. Other commonly observed outcomes of interest included pneumonia, ARDS, cardiovascular events, acute kidney injury and venous thromboembolic events. One report from the CDC published at the end of March 2020 reported that in the US, 21-31% of those tested were hospitalized^26^. In Europe, a report from the European Center for Disease Prevention and Control (ECDC) published in April 2020 reported that among all cases, 32% were hospitalized^27^. These proportions are likely to decline over time as less severe cases are increasingly tested with better testing capabilities. The fatality proportions reported in our study were slightly higher than the latest published data from the John Hopkins Coronavirus Resource Center (at 3% for both the US and Spain)^28^, largely because most of our data were from March to May 2020, with the outbreak at its peak. We also compared our results with the CDC report published at the end of May^29^, in which the reported percentage of deaths attributed to pneumonia, influenza or COVID-19 was 13.7%.

### Strengths and limitations

Our study has limitations. We analysed data collected during routine care and in actual practice settings for clinical rather than research purposes. It is therefore expected that some information will be incompletely reported, leading to potential misclassification. The variability observed in testing practices and the similarity in the distribution of important comorbidities, symptoms and demographics in the *tested* vs *tested*+ cohorts suggests targeted testing for subjects with severe forms of disease in the first months of the pandemic. The lack of mild infections limits our ability to identify key variables associated with susceptibility to infection. Underreporting of symptoms in EHR is another limitation of the study which can be due to factors such as source of data, testing availability, model of care, and reimbursement policies. We observed wide-spread variation in symptom reporting and a generally lower prevalence of symptoms compared to previous literature. Despite this, our study suggests that symptoms such as cough and fever remain key disease features, predictive of a positive test. We were not able to describe characteristics of the population tested negative for SARS-CoV-2 given the transient state of this population and specifically the difficulties in accurately ascertaining this cohort during a pandemic. However, the majority of the *tested* population consists of persons who tested negative for SARS-CoV-2 with *tested+* cohort comprising at most 6.7% of the *tested* cohort. As such, the observed difference in characteristics and outcomes between the *tested* and tested*+* cohorts are largely explained by the tested negative population.

This study also has strengths. First, this is the largest patient-level cohort of COVID-19 tested and *tested*+ patients to date. This allowed us to study the observed risks of relatively uncommon outcomes, otherwise not identifiable in smaller datasets. Secondly, we obtained data from multiple centres, three countries, three continents, and covering almost 6 months of 2020, allowing for a comprehensive characterisation of the study population, their key baseline characteristics, symptoms and 30-day outcomes. Finally, the use of routinely collected data from multiple sources maximized the external validity and generalizability of our findings to our settings internationally, whilst minimising Hawthorne effects and selection bias.

## Conclusions

In the largest cohort study to date, adult women were the majority of the almost 3.32 million people tested, and of the >219,000 tested positive study participants. Fever, cough and dyspnoea were the most common symptoms, more frequently reported in patients tested positive for SARS-CoV-2. The observed heterogeneity in testing practices complicated an accurate measurement of health outcomes related to COVID-19 in the first half of 2020. With increasing testing capacity, as recommended by the WHO^30^, the test+/tested ratio declined in May and June compared to March/April. Ensuring sufficient testing capacity will remain a challenge during the current second wave. If achieved, keeping a low (e.g. <5%) test+/tested ratio will help control and understand the COVID-19 pandemic in the coming months.

## Data Availability

Analyses were performed locally in compliance with all applicable data privacy laws. Although the underlying data is not readily available to be shared, authors contributing to this paper have direct access to the data sources used in this study. All results (e.g. aggregate statistics, not presented at a patient-level with redactions for minimum cell count) are available for public inquiry. These results are inclusive of site-identifiers by contributing data sources to enable interrogation of each contributing site. All analytic code and result sets are made available at: https://github.com/ohdsi-studies/Covid19CharacterizationCharybdis

https://github.com/ohdsi-studies/Covid19CharacterizationCharybdis

## ETHICAL CONSIDERATIONS

All the data partners received Institutional Review Board (IRB) approval or exemption. STARR-OMOP had approval from IRB Panel #8 (RB-53248) registered to Leland Stanford Junior University under the Stanford Human Research Protection Program (HRPP). The use of VA-OMOP data was reviewed by the Department of Veterans Affairs Central IRB, was determined to meet the criteria for exemption under Exemption Category 4(3) and approved for Waiver of HIPAA Authorization. The research was approved by the Columbia University Institutional Review Board as an OHDSI network study. The use of SIDIAP was approved by the Clinical Research Ethics Committee of the IDIAPJGol (project code: 20/070-PCV). The use of CPRD was approved by the Independent Scientific Advisory Committee (ISAC) (protocol number 20_059RA2). The use of IQVIA-OpenClaims was exempted from IRB approval. The CU-AMC study participation was considered exempt by the Colorado Multiple Institutional Review Board. The use of the UWM-CRD data was reviewed and approved by the University of Washington Institutional Review Board (STUDY 00010708).

## CONTRIBUTORSHIP STATEMENT

TDS, KK, APU, PR, DPA, FN, TMA, LMS, and AG conceived and designed the study. SLD, TF, KEL, MEM, KN, JDP, CGR, NHS, PR, KK, LMS, JAT, ABW and TDS coordinated data contributions at their respective sites. AP, AGS, TF, SFB, JDP, KK and TDS analyzed the data; AG and LL produced the figures and tables. AG, LL, DPA, JJ, FN, TMA. interpreted the data. AG, LL, DPA searched the literature and wrote the first draft. LL, AG, DV, CA, WA, PC, OA, JJ, VS, and HA reviewed the literature and revised the manuscript. All authors contributed to the revision of the first draft, reviewed and approved the final version of the manuscript.

## ACKNOWLEDGEMENTS

We would like to acknowledge the patients who suffered from or died of this devastating disease, and their families and carers. We would also like to thank the healthcare professionals involved in the management of COVID-19 during these challenging times, from primary care to intensive care units.

