## Supplementary Materials for "Baseline phenotype and 30-day outcomes of people tested for COVID-19: an international network cohort including >3.32 million people tested with real-time PCR and >219,000 tested positive for SARS-CoV-2 in South Korea, Spain and the United States"

### Supplementary Table 1. Contributing databases to OHDSI's open science community efforts to characterize the natural history of COVID-19

| Location | Full Database Name | Type of data | Included in the study | Point of contact, name (email address) |
|---|---|---|---|---|
| South Korea | Health Insurance Review & Assessment Service | National claims | Yes | Seng Chan You: |
| Spain | Information System for Research in Primary Care | Primary Care-Only EHR | Yes | Talita Duarte Salles:<br>Sergio Fernandez Bertolin: |
| US | Columbia University Irving Medical Center | Hospital EHR | Yes | George Hripcsak:<br>Karthik Natarajan:<br>Thomas Falconer:<br>Matthew Spotnitz:<br>Anna Ostropolets: |
| US | HealthVerity | National claims | Yes | Clair Blacketer:<br>Frank DeFalco: |
| US | IQVIA Open Claims | National claims | Yes | Christian Reich:<br>Kristin Kostka: |
| US | Premier | Hospital EHR | Yes | Alan Andryc:<br>Stephen Fortin: |
| US | Tufts Research Data Warehouse | Hospital EHR | Yes | Andrew Williams: |
| US | United States Department of Veterans Affairs | Hospital EHR | Yes | Scott L. Duvall:<br>Kristine Lynch:<br>Michael Matheny: |
| US | U of Colorado Anschuz Medical Campus Health Data Compass | Hospital EHR | Yes | Lisa M. Schilling:<br>William Carter: |
| US | Optum® de-identified COVID-19 Electronic Health Record dataset | Hospital EHR | Yes | Clair Blacketer:<br>Frank DeFalco: |
| US | UW Medicine COVID Research Dataset | Hospital EHR | Yes | Jason A. Thomas:<br>Adam B. Wilcox: |
| Germany | IQVIA Disease Analyser Germany | Primary Care-Only EHR | No | NA |
| South Korea | Daegu Catholic University Medical Center | Hospital EHR | No | NA |
| Spain | HM Hospitals | Hospital EHR | No | NA |
| The Netherlands | Integrated Primary Care Information | Primary Care-Only EHR | No | NA |
| France | IQVIA Longitudinal Patient Data (LPD) France | Primary Care-Only EHR | No | NA |
| China | Nanfang Hospital COVID-19 Research Database (NFHCRD) | Hospital EHR | No | NA |
| Spain | Information System for Research in Primary Care Hospitalization Linked Data (SIDIAP-H) | Primary Care-Only EHR linked to hospital records | No | NA |
| UK | Clinical Practice Research Datalink | Primary Care-Only EHR | No | NA |
| US | Optum® Socio-Economic Status (SES) | Hospital EHR | No | NA |

*EHR: electronic health record

### Supplementary Table 2. Description of databases included in the study

| Country | Name | Description |
|---|---|---|
| US | HealthVerity | This HealthVerity derived data set contains de-identified patient information with an antibody and/or diagnostic test for COVID-19 linked to all available Medical Claims and Pharmacy Data from select private data providers participating in the HealthVerity marketplace. |
| US | Premier | Complete clinical coding, hospital cost, and patient billing data from approximately 700 hospitals throughout the United States representing 20% of inpatient hospital stays. |
| US | STARR-OMOP | A clinical data warehouse containing live Epic data from Stanford Health Care, the Stanford Children's Hospital, the University Healthcare Alliance and Packard Children's Health Alliance clinics and other auxiliary data from Hospital applications such as radiology PACS. https://arxiv.org/abs/2003.10534 |
| US | Tufts Research Data Warehouse (TRDW) | Electronic medical record data on approximately 1 million patients who received care beginning in 2006 at Tufts Medical Center (TMC). |
| US | Department of Veterans Affairs | VA OMOP data reflects the national Department of Veterans Affairs health care system, which is the largest integrated provider of medical and mental health services in the United States. Care is provided at 170 VA Medical Centers and 1,063 outpatient sites serving more than 9 million enrolled Veterans each year. |
| US | IQVIA Open Claims | Pre-adjudicated claims covering over 300 Million lives (~80% of the US) collected from office-based physicians and specialists via office management software and clearinghouse switch sources for the purpose of reimbursement |
| US | Columbia University Irving Medical Center | The clinical data warehouse of New York-Presbyterian Hospital/Columbia University Irving Medical Center, New York, NY, based on its current and previous electronic health record systems, with data spanning over 30 years and including over 6 million patients |
| US | U of Colorado Anschuz Medical Campus Health Data Compass | |
| US | Optum EHR | Optum© de-identified Electronic Health Record Dataset represents Humedica's Electronic Health Record data a medical records database |
| US | UW Medicine COVID Research Dataset | The dataset includes UW Medicine patients who have been tested for COVID-19 that have information within the UW Medicine Electronic Health Record. The dataset is a subgroup of the non-OMOP clinical data warehouse of University of Washington Medical Center, comprised of Harborview Medical Center, UW Medical Center - Montlake, and Northwest hospital in Seattle WA, and is based on its current electronic health record systems, with data spanning over 10 years and including roughly 5 million patients |
| Spain | Information System for Research in Primary Care (SIDIAP) | The Information System for Research in Primary Care (SIDIAP; www.sidiap.org) is a primary care records database that covers approximately 7 million people, equivalent to an 80% of the population of Catalonia, North-East Spain. Healthcare is universal and tax-payer funded in the region, and primary care physicians are gatekeepers for all care and responsible for repeat prescriptions. |
| South Korea | Health Insurance Review & Assessment Service | National claim data from a single insurance service from South Korea |

### Supplementary Table 3. Definitions and codes used to identify persons tested for SARS-CoV-2 and patients tested positive for SARS-CoV-2

The below tables summarize the concepts used to identify patients tested for SARS-CoV-2 (*tested* cohort) and patients tested positive for SARS-CoV2 (*tested+* cohort). The full description of the logic used is provided at https://atlas.ohdsi.org/#/cohortdefinition/206 for *tested* and https://atlas.ohdsi.org/#/cohortdefinition/204 for *tested+* cohorts.

**COVID-19 testing - Positive**

| Id | Name | Vocabulary |
|---|---|---|
| 756055 | Measurement of Severe acute respiratory syndrome coronavirus 2 (SARS-CoV-2) | OMOP Extension |

**COVID-19 specific test Positive**

| Id | Name | Vocabulary |
|---|---|---|
| 37310282 | 2019 novel coronavirus detected | SNOMED |
| 37310281 | 2019 novel coronavirus not detected* | SNOMED |
| 756055 | Measurement of Severe acute respiratory syndrome coronavirus 2 (SARS-CoV-2) | OMOP Extension |

*used to only include measurements with a value equal to detected, positive or present

### Supplementary Figure 1. Database selection process flowchart

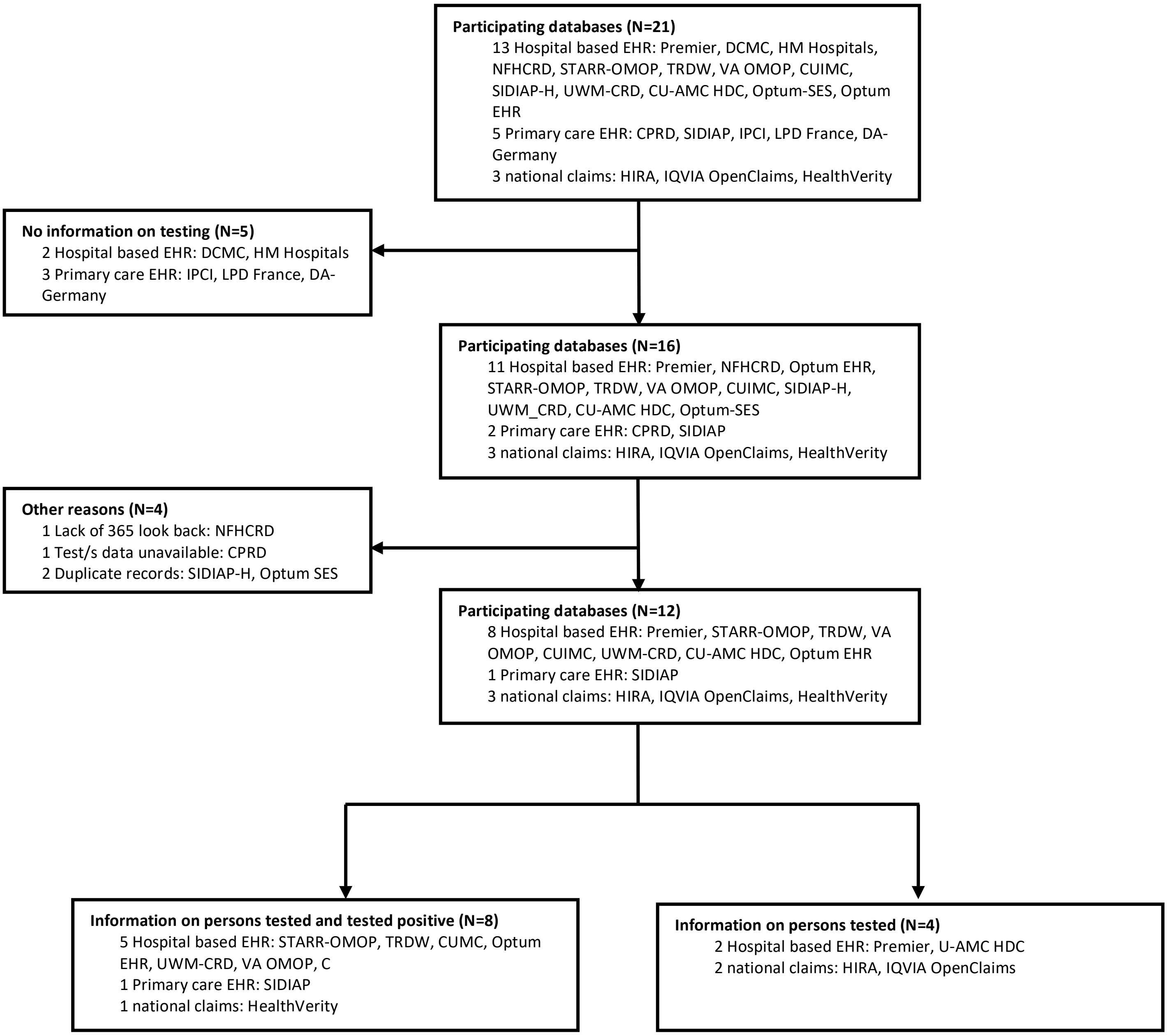


Clinical Practice Research Datalink (CPRD), U of Colorado Anschuz Medical Campus Health Data Compass (CU-AMC HDC), Columbia University Irving Medical Center (CUIMC), Disease Analyzer (DA) Germany, Daegu Catholic University Medical Center (DCMC), Health Insurance Review & Assessment Service (HIRA), Hospital de Madrid (HM) Hospitals, Integrated Primary Care Information (IPCI), longitudinal Patients Database ( LPD ) France, Nanfang Hospital COVID-19 Research Database (NFHCRD), Optum® de-identified COVID-19 Electronic Health Record dataset (Optum EHR), Optum Socio-Economic Status (OPTUM-SES), Information System for Research in Primary Care (SIDIAP), SIDIAP– Hospitalization Linked Data (SIDIAP-H), Tufts Research Data Warehouse (TRDW), UW Medicine COVID Research Dataset (UWM-CRP), Department of Veterans Affairs (VA-OMOP)

**Supplementary Figure 2: Baseline comorbidities up to 30 days prior to first test or first positive test, respectively, among SARS-CoV-2 *tested* and *tested+* cohorts across databases of various setting, stratified by (a) sex and (b) in persons <18 (paediatrics) and > 65 years old (elderly)**

**2a.**

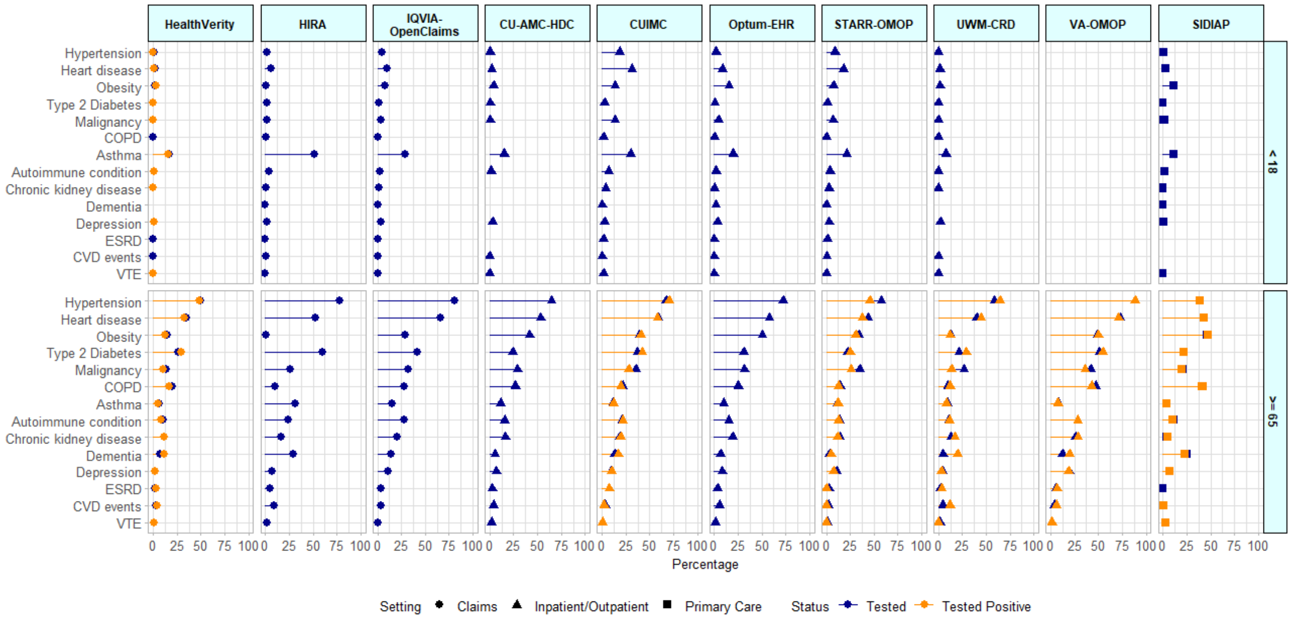


**2b.**

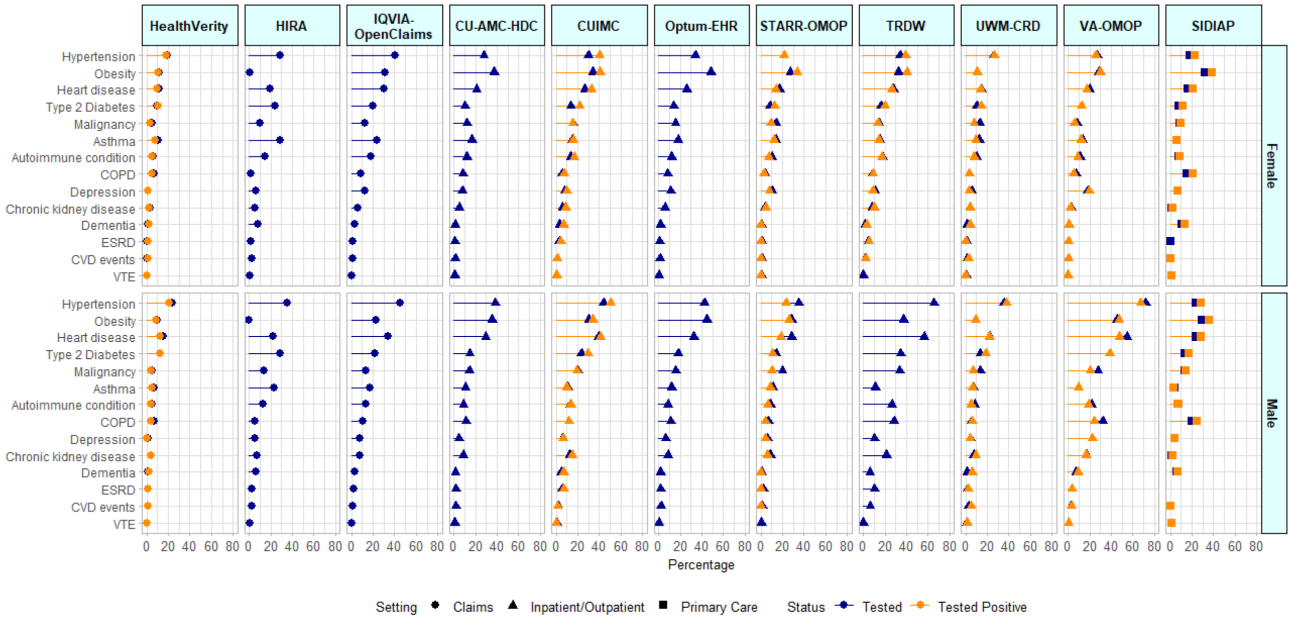


*COPD = Chronic obstructive pulmonary disease; ESRD = End stage renal disease; CVD = Cardiovascular disease; VTE = Venous thromboembolism events*

**Supplementary Figure 3: COVID-19 symptoms at index date among SARS-CoV-2 *tested* and *tested+* cohorts across databases of various setting, (a) stratified by sex, (b) in persons <18 (pediatrics) and > 65 years old (elderly)**

**3a.**

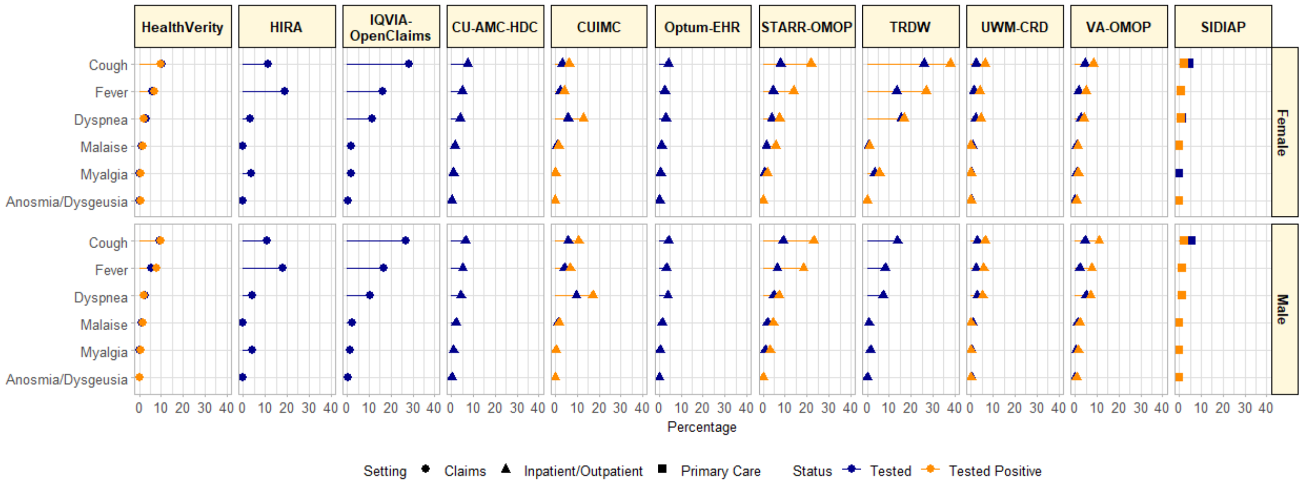


**3b.**

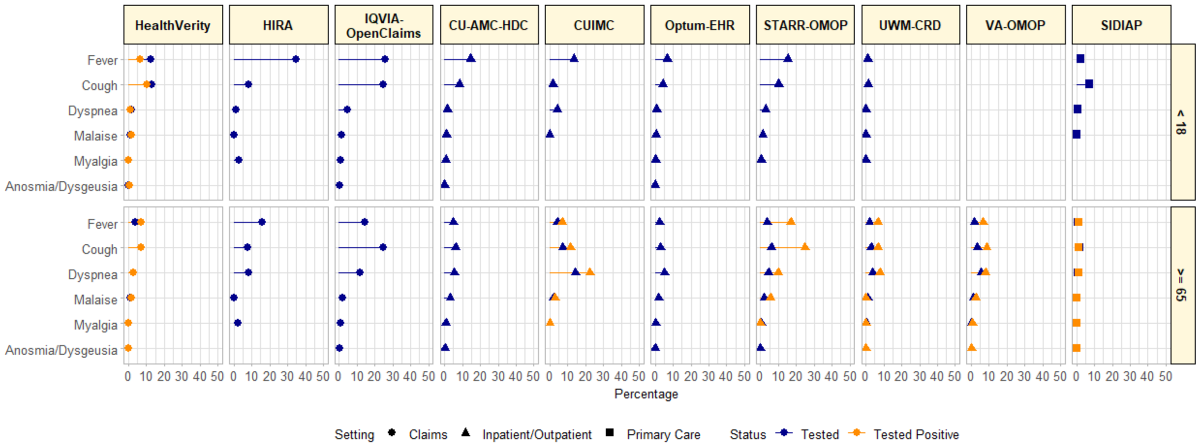

**Supplementary Figure 4: All clinical outcomes during hospitalization one-month post index date among SARS-CoV-2 *tested+* cohorts across databases of various setting**

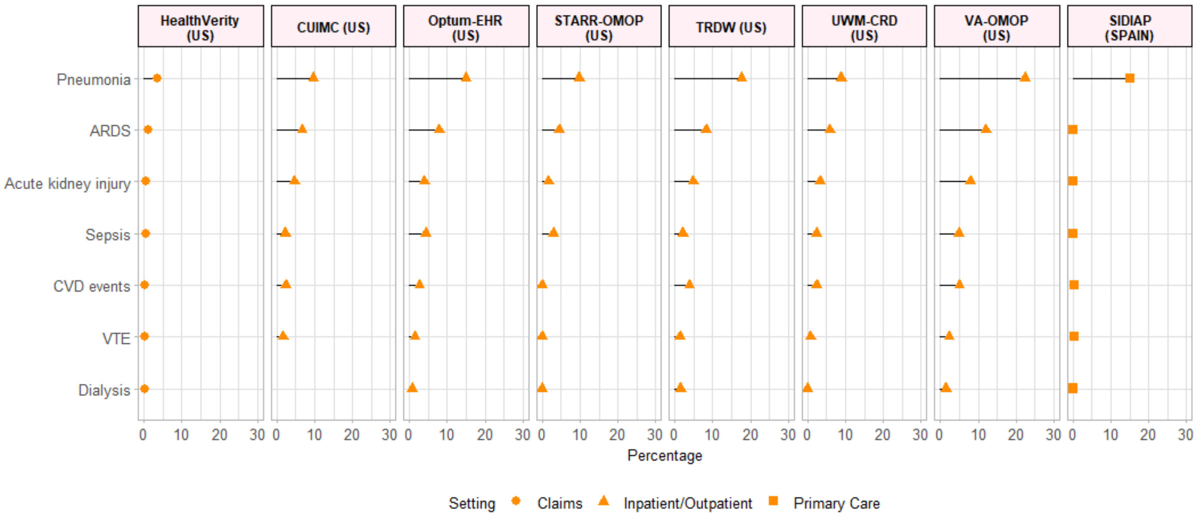


*ARDS = Acute respiratory distress syndrome; CVD = Cardiovascular disease; VTE = Venous thromboembolism events*
